# Combined omic analyses reveal novel loss-of-function *NLGN3* variants in GnRH deficiency and autism

**DOI:** 10.1101/2022.05.24.22275221

**Authors:** Roberto Oleari, Antonella Lettieri, Stefano Manzini, Alyssa Paganoni, Valentina André, Paolo Grazioli, Marco Busnelli, Paolo Duminuco, Antonio Vitobello, Christophe Philippe, Varoona Bizaoui, Helen L. Storr, Federica Amoruso, Fani Memi, Valeria Vezzoli, Valentina Massa, Peter Scheiffele, Sasha R. Howard, Anna Cariboni

**Affiliations:** Department of Pharmacological and Biomolecular Sciences, University of Milan, Milan, Italy; CRC Aldo Ravelli for Neurotechnology and Experimental Brain Therapeutics, Department of Health Sciences, University of Milan, Milan, Italy; Department of Health Sciences, University of Milan, Milan, Italy; Laboratory of Endocrine and Metabolic Research, IRCCS Istituto Auxologico Italiano, Cusano Milanino, Italy; Unité Fonctionnelle Innovation en Diagnostic Génomique des Maladies Rares, FHU-TRANSLAD, CHU Dijon Bourgogne, Dijon, France; INSERM UMR 1231 GAD (Génétique des Anomalies du Développement), Université de Bourgogne, Dijon, France; Genetics and neurodevelopment Centre Hospitalier de l’Estran, Pontorson, France; Centre for Endocrinology William Harvey Research Institute Barts and the London School of Medicine and Dentistry, Queen Mary University of London, London, United Kingdom; Royal London Children’s Hospital, Barts Health NHS Trust, London, United Kingdom; Wellcome-MRC Cambridge Stem Cell Institute, Jeffrey Cheah Biomedical Centre, Cambridge, United Kingdom; Biozentrum of the University of Basel, Basel, Switzerland

**Author notes:** Equally contributed (Roberto Oleari, Antonella Lettieri).

**Keywords:** GnRH neurons, transcriptome, NLGN3, delayed puberty, autism spectrum disorder.

## Abstract

Gonadotropin releasing hormone (GnRH) deficiency is a disorder characterized by absent or delayed puberty, with largely unknown genetic causes. The purpose of this study was to obtain and exploit gene expression profiles of GnRH neurons during development to unveil novel biological mechanisms and genetic determinants underlying GnRH deficiency (GD). Here, we combined bioinformatic analyses of primary embryonic and immortalized GnRH neuron transcriptomes with exome sequencing from GD patients to identify candidate causative genes. Among differentially expressed and filtered transcripts, we found loss-of-function (LoF) variants of the autism-linked *Neuroligin 3* (*NLGN3*) gene in two unrelated patients co-presenting with GD and neurodevelopmental traits. We demonstrated that *NLGN3* is upregulated in maturing GnRH neurons and that NLGN3 wild type, but not mutant proteins, promotes neuritogenesis when overexpressed in developing GnRH cells. Our data represent proof-of-principle that this complementary approach can identify novel candidate GD genes and demonstrate that LoF *NLGN3* variants may contribute to GD. This novel genotype-phenotype correlation implies common genetic mechanisms underlying neurodevelopmental disorders, such as GD and autistic spectrum disorder.

## Introduction

Gonadotropin-releasing hormone (GnRH) is the master hormone regulating the hypothalamic-pituitary-gonadal (HPG) reproductive axis and its pulsatile secretion is crucial for puberty onset, sexual maturation and fertility (1). GnRH is produced by a small number of hypothalamic neuroendocrine neurons called GnRH neurons which, during embryonic development, originate in the nasal placode (NP) and migrate along vomeronasal nerves to reach the hypothalamus (2).

Disruption in GnRH neuron development or hypothalamic function are the leading causes of genetic reproductive disorders such as Hypogonadotropic Hypogonadism (HH) and Kallmann Syndrome (KS), which are characterized by absent or delayed puberty, due to GnRH deficiency (GD) (3). The known genes causing GD account for only 50% of cases (3, 4), supporting the need for studying the genetic signatures of GnRH neurons as a novel strategy to expedite etiological discovery in the remaining cases. However, the study of the GnRH neuronal system is hampered by the difficulty in obtaining primary GnRH neurons, which are small in number and lack specific markers. To overcome these issues, alternative tools, such as immortalized GnRH neuron cell lines (5) and reporter rodent and zebrafish lines (6–8), have been adopted. Recently, RNA sequencing has been applied to obtain the transcriptome of iPSC-derived human GnRH neuron progenitors and early postmitotic GnRH neurons (9). Yet, no reports of gene expression profiles of primary mouse GnRH neurons during the whole developmental process are available.

Here we obtained for the first time the transcriptomic profiles of primary and immortalized embryonic GnRH neurons. Further, by combining filtering strategies with exome sequencing from human patients and *in vitro* functional experiments, we identified *Neuroligin 3* (*NLGN3*) as a new GD-disease candidate gene. *NLGN3* belongs to the neuroligin family, a class of postsynaptic cell adhesion molecules which regulate synapse organization (10) and dendritic outgrowth (11), and *NLGN3* missense variants have been so far associated with autistic spectrum disorder (ASD) (12). Thus, in this work we described two patients carrying novel nonsense *NLGN3* variants and presenting with clinical features typical of GD and ASD, therefore providing the first genetic correlation between these two neurodevelopmental disorders, which was previously only implied by a registry-based association study (4).

## Results

### The analysis of rat Gnrh1-GFP neuron transcriptome at three developmental stages revealed age-specific expression signatures

We first obtained the transcriptomic profiles of primary GnRH neurons isolated from pooled *Gnrh1*-GFP rat embryos (7) at embryonic day (E) 14, E17 and E20 by using Affymetrix GeneChip arrays (n = 1 per each stage; GSE174896; **Figure 1A**). Sample clustering by principal component analysis (PCA) investigation showed that genotype (i.e. GFP expression) rather than developmental stage mainly affects gene expression, causing GFP^-^ cells (E17 and E20) to cluster away from the GFP^+^ populations, except for the earlier stage (E14) (**Figure 1B**). Conversely, amongst GFP^+^ cells, the developmental stage strongly impacted on gene expression with samples clustering away one from each other (**Figure 1B**). By examining GFP^+^ cell transcriptomes, we identified expression changes in 4,844 detected probes, mapping to 2,739 unique annotated genes, using the E14 stage as common baseline (log_FC_ > 1). Specifically, we found 770 genes with an expression difference in excess of 2 fold change between E14 and E17, 1062 genes between E14 and E20, while 907 genes had at least a 2 fold change difference in expression in both conditions compared to E14, highlighting that each developmental stage is defined by a unique transcriptomic signature (**Figure 1E**). Among these stage-specific genes, we identified genes previously implicated in GnRH neuron development by direct experimental evidence (e.g. *Sirt1*, *Reln*, *Cxcr7*, *Slit2*, *Lif*) (13, 14) or found mutated in GD patients (e.g. *Sema3a*, *Sema7a*, *Fgfr1*, *Nrp1*) (3).

**Figure 1.**
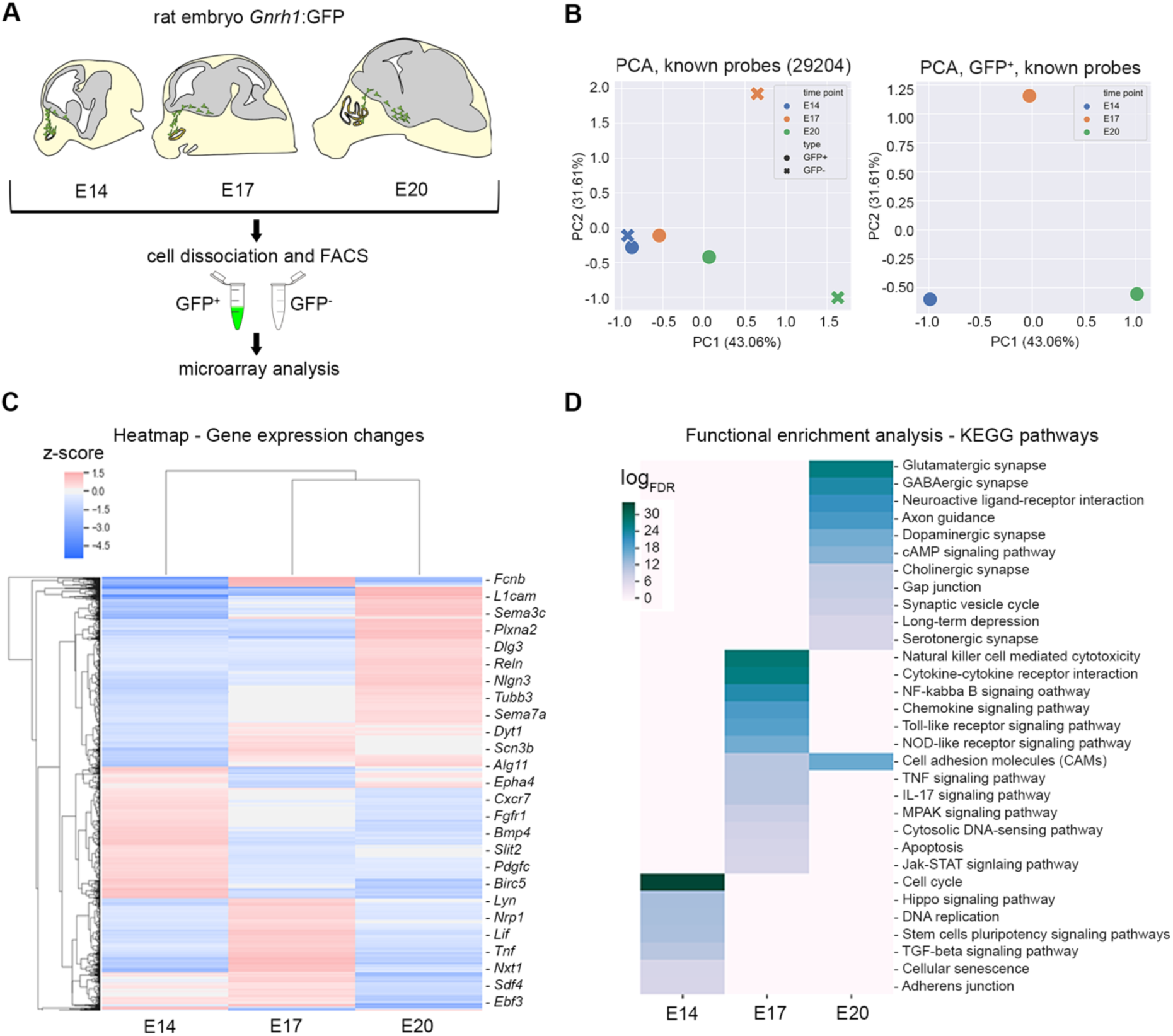
Transcriptomic analysis of GnRH neurons from *Gnrh1*-GFP rats at different developmental time points. A - Schematic drawing representing *Gnrh1*-GFP rat embryos at indicated developmental time points and localization of GnRH neurons (in green). GFP^+^ cells were isolated after embryo dissociation with FACS and RNA from GFP^+^ cells were used for microarray analysis. B - Dimensionality reduction was performed on gene expression space for each sample, and the first 2 principal coordinates are charted. Samples are shown for each time point (E14 - blue, E17 - orange, E20 - green). Circles represent GFP^+^ cells, crosses GFP^-^ cells. C - The ratio of the log_2_ expression value and mean value across all samples for each probe has been calculated and clustered hierarchically (Euclidean distance metric). Red indicates higher expression, blue lower expression. The heatmap reveals three broad gene clusters. D - Functional enrichment analysis has been carried out for genes belonging to previously identified clusters. Enriched KEGG pathways (FDR < 0.01) are summarized in an heatmap for genes upregulated at E14, E17 and E20 timepoints. Enrichment scores are reported as log_FDR_; higher values (deep blue) indicate highly enriched pathways, lower values (grey) indicate poorly enriched pathways. Abbreviations: NP, nasal placode; HYP, hypothalamus.

Accordingly, functional enrichment analysis, performed with reString software (15) and based on KEGG pathway annotations, revealed significantly enriched pathways (FDR < 0.01) that are consistent with those activated along the developmental stage (**Figure 1D**). In rat embryos, GnRH neurons are mainly located in the NP at E14, reach the peak of migration at E17 and cease to migrate by E20. Accordingly, we found that highly represented pathways contained: genes commonly active during early NP patterning at E14 (e.g. *Bmp4*, *Fgfr1*, *Gli2*; **Supplemental figure 1A**) (14, 16); genes associated with response to chemokine and cytokine-receptors interactions at E17 (e.g. *Csf1*, *Csf1r*, *Lif*; **Supplemental figure 1B**) (14); genes related to axon growth and synapse function at E20 (e.g. *Sema6b*, *Plxna2/4*, *Sema7a*, *Robo2*, *Slit1*, *Epha5*, *L1cam*) (17) (**Supplemental figure 1C**). Similar results were obtained with the functional enrichment analysis of Gene Ontology (GO) Biological Processes (BP) (FDR < 10^-5^; **Supplemental figure 1D**).

Overall, a high number of stage-specific genes enriched biological pathways relevant to and consistent with the different phases of GnRH neuron development, such as differentiation and migration at early phases (E14, E17) and establishment of neuron projections at later phases (E20) (14, 16).

### The combined data mining of primary and immortalized GnRH neuron transcriptomes with GD causative genes prioritized 20 candidate genes

To refine the list of candidate genes, we leveraged an integrated approach based on functional enrichment analysis, transcriptomic profiles of immortalized GnRH neurons and prioritization bioinformatic tools (**Figure 2**). We first selected genes belonging to significantly enriched pathways related to cytoskeleton dynamics, migration and axon development for each time point (FDR < 0.01 for KEGG pathways and GO BP; **Supplemental table 1 and 2**).

**Figure 2.**
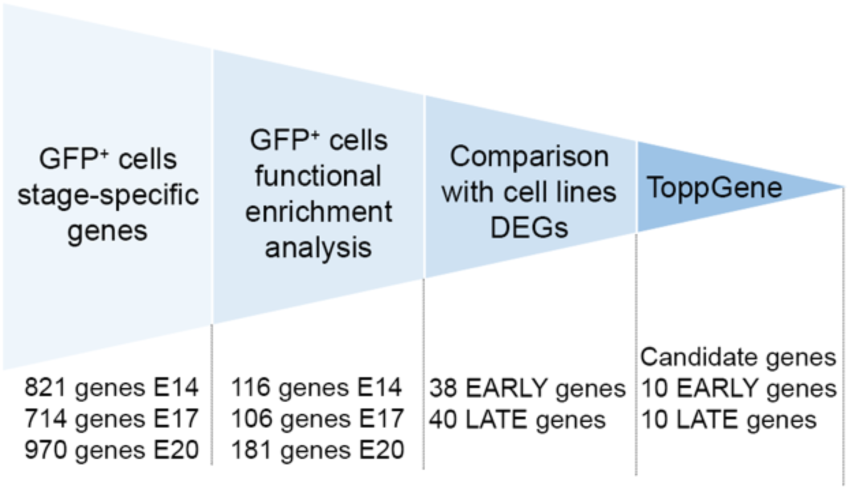
Filtering strategy to reveal novel candidate genes underlying GD pathogenesis. Flowchart of filtering strategy applied to narrow down the list of candidate genes. Upregulated genes at each time point were filtered upon selection of GO/KEGG pathways related to cytoskeleton dynamics, migration and axon development (see Table S1 for the full list of pathways). Selected genes were crossed with DEGs from GN11 and GT1-7 cell lines belonging to the same pathways, delineating two groups of genes, named “early” and “late” genes. Last, candidate genes were prioritized with ToppGene software and top10 output genes from both groups were used for subsequent analyses.

Owing to technical constraints, RNA from primary neurons was pooled and run as a single replicate. Because of this, we next aimed to validate these results using datasets from immortalized neurons. The microarray analysis data was integrated with DEGs obtained from GN11 and GT1-7 cells (n = 3; GSE174902; **Figure 3A**), which are established models of immature migrating and maturing mouse GnRH neurons, respectively (5). PCA analysis revealed a high similarity within samples of each cell line (**Figure 3B**) and 1,815 differentially expressed probes (adjusted *P* value < 0.05, log_FC_ > 2), mapping to 1,546 unique annotated genes (**Figure 3C,D**), were found. Functional enrichment analysis (**Figure 3E and Supplemental figure 2A**) showed that genes up-regulated in GN11 cells significantly populated pathways related to migration and progenitor maintenance (**Supplemental figure 2B**). Conversely, the pathways most represented in GT1-7 cells belonged to neuronal maturation processes (e.g axon development and synapse assembly) (**Supplemental figure 2C**). The identified signatures from cell lines had a high level of overlap with the ones observed in primary neurons (**Figure 4A**). We therefore integrated these two datasets to obtain two gene set clusters that we named ‘early’ and ‘late’ genes. Among them, we considered only genes that enriched selected pathways shared by both GFP^+^ primary neuronal cells and immortalized GnRH neurons.

**Figure 3.**
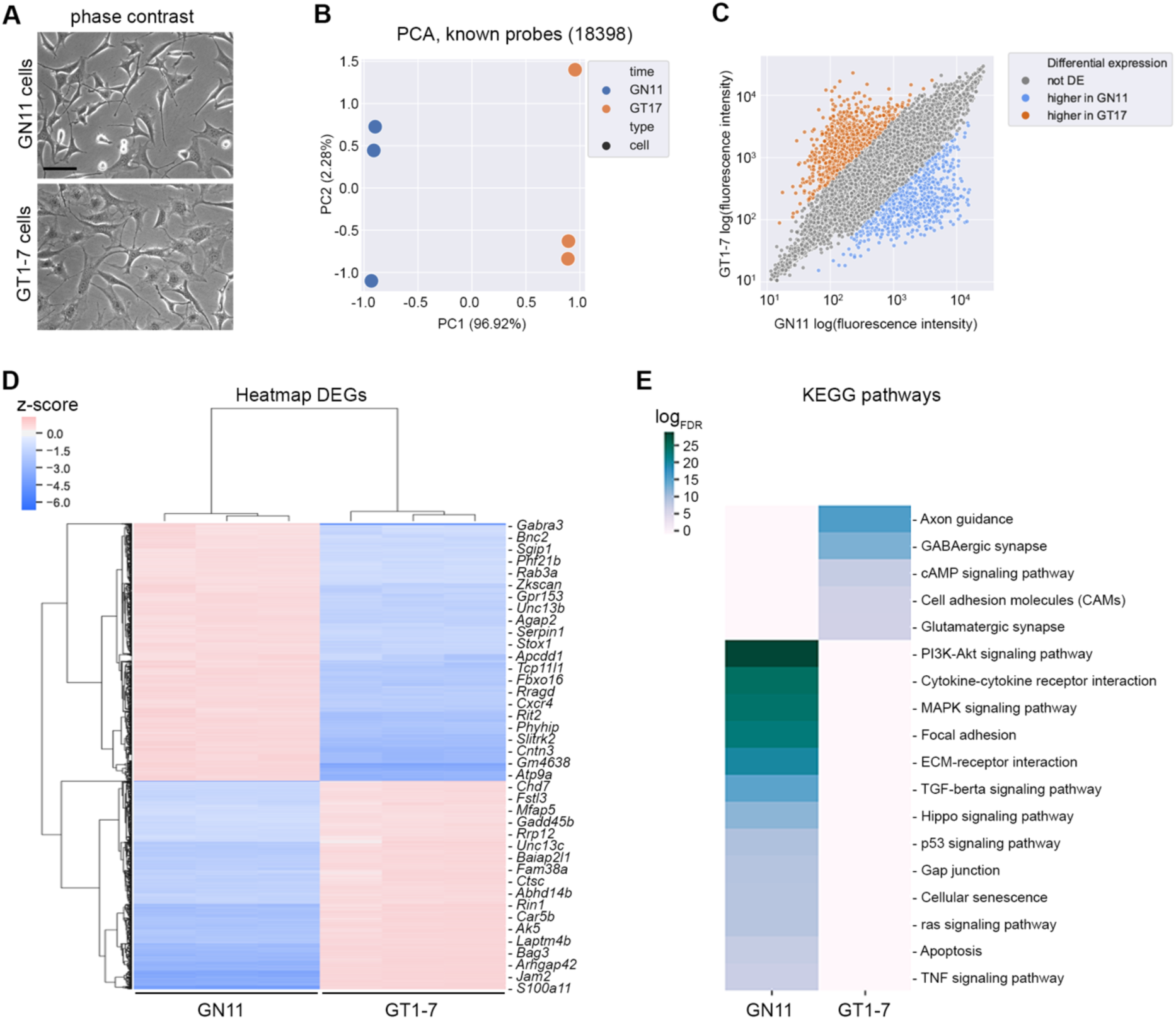
Transcriptomic analysis of GN11 and GT1-7 immortalized GnRH neurons. A - Phase contrast microphotographs of GN11 and GT1-7 cells. B - Dimensionality reduction was performed on gene expression space (annotated probes only) for each GN11 and GT1-7 cell sample, and the first 2 principal coordinates are charted. C - Scatter plot showing DEGs (log_FC >_ 2, adjusted *P* value < 0.05) between GN11 and GT1-7 cells. GN11 upregulated genes are indicate in blue whereas GT1-7 upregulated genes are in orange; grey dots indicate non DEGs. D - The ratio of the log_2_ expression value and mean value across all samples for each probe has been calculated and clustered hierarchically (Euclidean distance metric) for DEGs (log_FC >_ 2) between GN11 and GT1-7 cells. Red indicate higher expression, blue lower expression. E - Enriched KEGG pathways (E, FDR < 0.01) found by STRING functional enrichment analysis computed on DEGs between GN11 and GT1-7 cells. Enrichment scores are reported as log_FDR_; higher values (deep blue) indicate highly enriched pathways, lower values (grey) indicate poorly enriched pathways.

**Figure 4.**
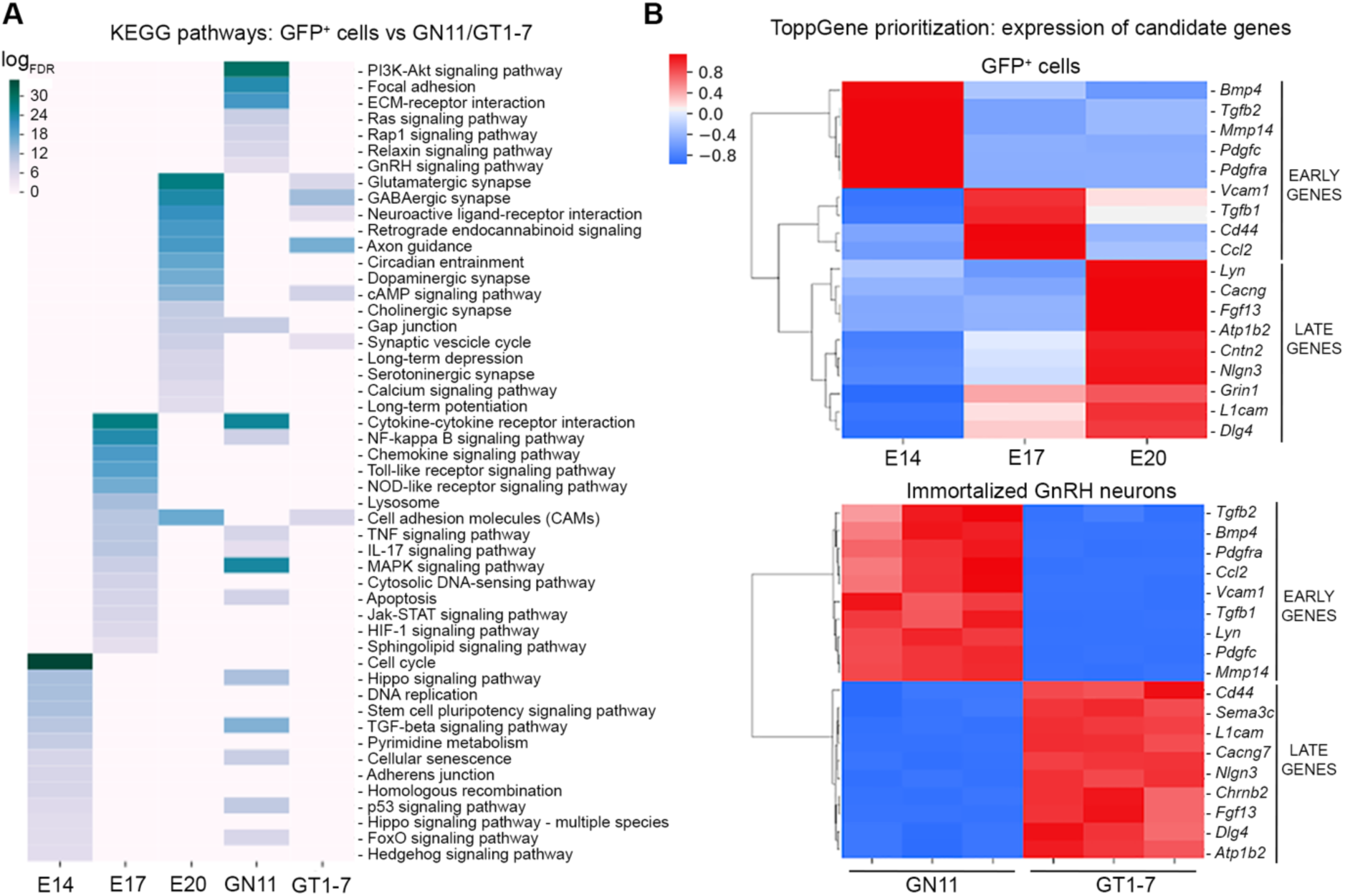
Filtering analyses revealed 20 candidate genes that differently enriched early and late stages of development. A - The functional enrichment analysis detailed in Figure 1F is integrated with the DEG genes in GN11 vs GT1-7 immortalized cells lines. Genes upregulated in GN11 cells vs GT1-7 enrich in the same pathways (FDR < 0.01) as GFP^+^ cells at E14 and E17. Notably, a specific subset of enriched pathways specific to GN11 cells exists. Conversely, genes upregulated in GT1-7 cells mostly enriched in the same pathways as GFP^+^ cells at E20. Enrichment scores are reported as log_FDR_; higher values (deep blue) indicate highly enriched pathways, lower values (grey) indicate poorly enriched pathways. B - Z-scored gene expression values of Top10 output early and late genes after ToppGene prioritization in GFP^+^ and immortalized cells along with embryonic development.

Last, to identify among filtered genes those that could be potentially implicated in GD pathogenesis, ToppGene software (18) was applied to rank candidates based on functional or structural similarity with a set of known GD causative ‘input’ genes (3, 19) (**Supplemental table 3**). The top ten output genes for each list (early or late) were selected (**Figure 4B and Table 1**) and the expression trends of two representative early and late genes were validated in GN11 and GT1-7 cells (**Supplemental figure 3**).

**Table 1.**
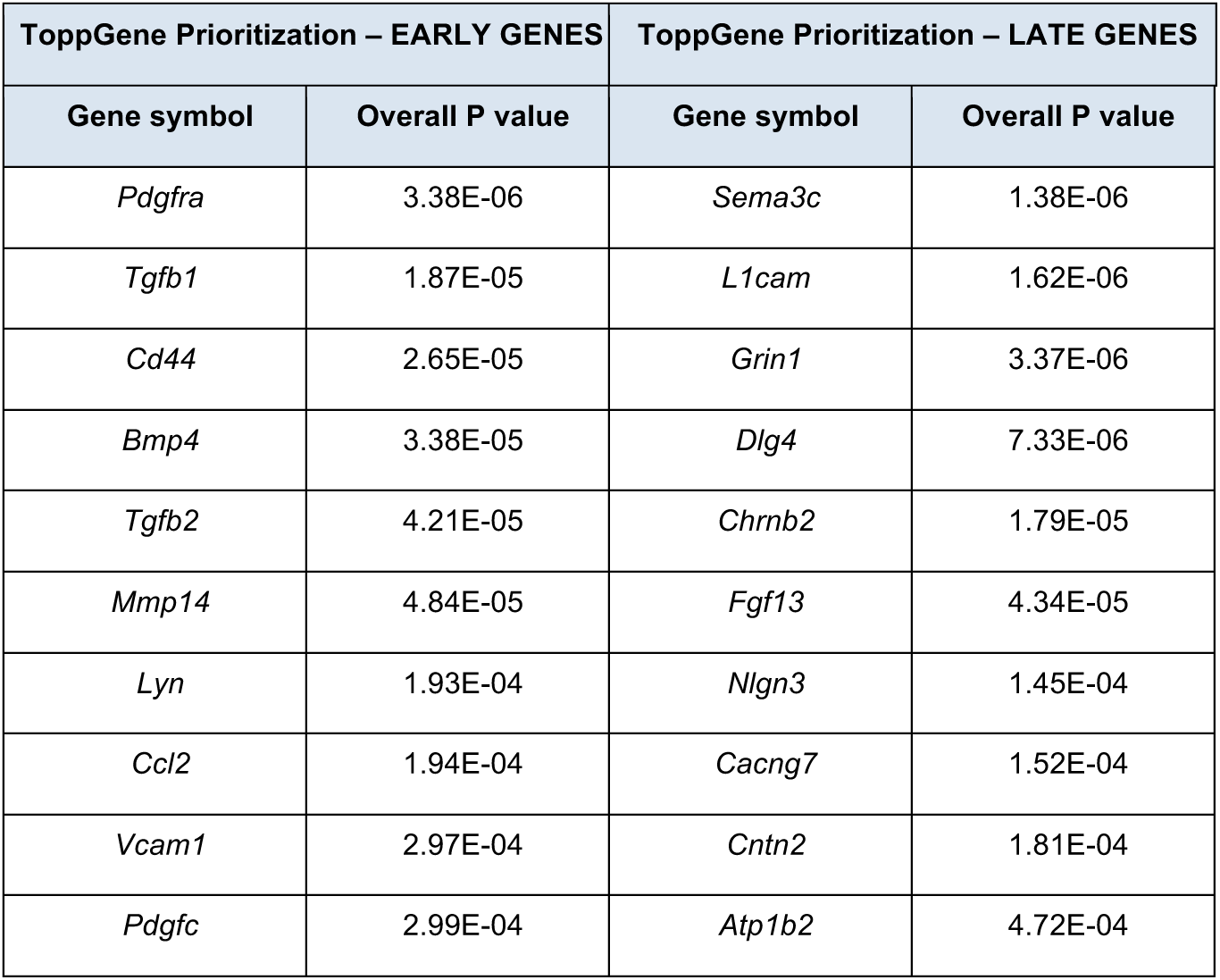
List of the top 10 output early and late genes after ToppGene prioritization with relative overall P values. ToppGene was instructed with the known GD genes (see Supplemental Table 3 for the list of genes) to prioritize genes and the top 10 output genes were considered for further analyses.

In summary, the microarray analyses combined with candidate gene filtering helped to identify 20 genes (from now on named “top 20 genes”) that have not been previously linked with GnRH neuron biology or GD.

### NLGN3 candidate gene is mutated in two patients affected by GD and ASD

To validate the relevance of the top 20 genes in GD pathogenesis, the presence of potentially deleterious variants for each gene was interrogated in exomes from our cohort of GD patients (n = 47) (20). A male patient (Case 1) with partial GD (with puberty that had initiated and then arrested (21)) and ASD traits was found to carry a predicted loss-of-function (LoF) variant in the X-linked *NLGN3* gene (Gene ID 5441; **Figure 5A**). By interrogating GeneMatcher (22), we found an additional *NLGN3* LoF variant in an unrelated patient with phenotypic features of GD and ASD with intellectual disability (Case 2; **Figure 5A**).

**Figure 5.**
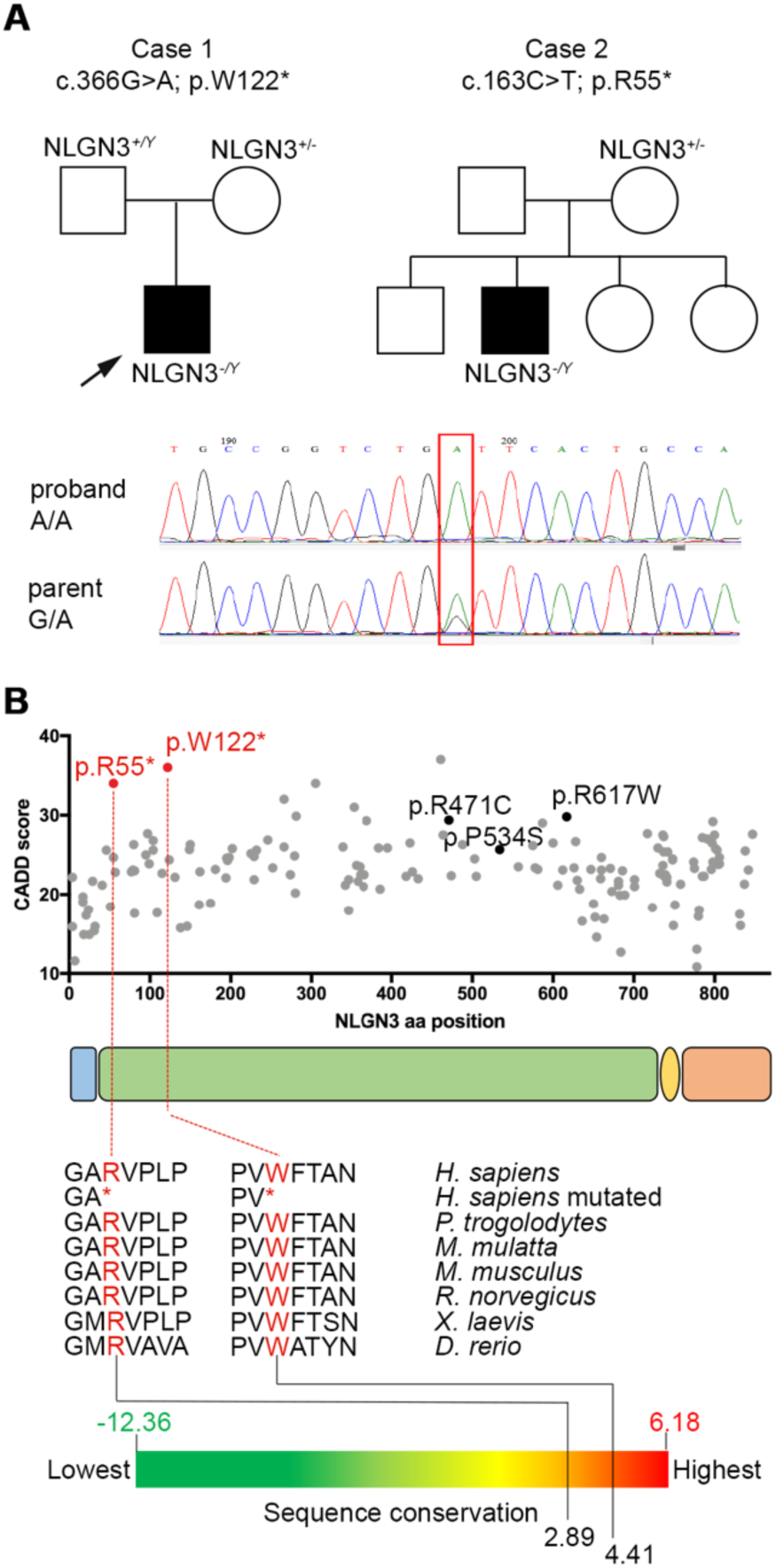
Two novel *NLGN3* nonsense variants found in patients with GD and ASD. A - Family pedigrees of the two probands (case 1 and case 2) identified in this study and presenting GD and ASD features. Sequence chromatograms of nucleotides 70367955-70367975 of the *NLGN3* coding sequence in proband 1 and his unaffected parent (the position of the G>A is highlighted by the red box). B - Schematic representation of NLGN3 protein (encoded by the NM_181303.1 transcript): signal peptide (sp, light blue), extracellular (green), transmembrane (tm, yellow) and intracellular (red) domains. All missense variants (grey dots) reported in gnomAD database (v2.1.1) are plotted according to their aminoacidic position and CADD score. Pathogenic variants related to ASD are indicated in black: R471C (29), P534S (26) and R617W (30). Identified variants (R55* and W122*) are indicated in red and have a higher CADD score compared to others. Multi-species alignment of partial protein sequences of vertebrate NLGN3 orthologue proteins shows that the R55 and W122 residues are evolutionarily conserved in humans and other vertebrate species with a high conservation degree, calculated by GERP++.

Case 1 was found to be hemizygous for a stop-gain variant in *NLGN3* (NM_181303.1: c.366G>A, p.W122*) (**Figure 5A,B).** He was born from healthy non-consanguineous parents, and his parent was found to be heterozygous for the same variant, as confirmed by Sanger sequencing (**Figure 5A**). Clinically, the proband presented in early adulthood with a picture of pubertal arrest with biochemical evidence of GD (**Table 2**). The proband reported late onset of puberty and had achieved testes volume of 10 mL by the age of first review. He was commenced on testosterone esters which were gradually increased up to a maximal dose of 250 mg every 4 weeks, but testes volume had increased only to 12 mL at age of more than 20 years, consistent with partial GD phenotype (3). In addition, he had associated phenotypic features including obesity (BMI = 36, height 178.5 cm), depression, anxiety, and social and communication difficulties consistent with ASD. Inhibin B concentrations were in the low-normal range but mildly elevated TSH concentrations with low-normal free T4 were reported, likely secondary to obesity (**Table 2**). MRI scan showed a normal appearance of the pituitary gland, pituitary stalk and hypothalamus (data not shown). There was no family history of GD, but his parent, who carried the same variant in *NLGN3*, was also obese.

**Table 2.** Case 1 and case 2 probands clinical data. Case 1 showed incomplete pubertal development in early adulthood with biochemical evidence of low serum testosterone and low basal gonadotropins. Whilst on treatment with testosterone esters his serum testosterone increased into the low-normal range, but testes volumes increased only to 12 mL, consistent with GD phenotype. Inhibin B concentrations were in the low-normal range with mildly elevated TSH concentrations with low-normal free T4 likely secondary to obesity (BMI 36). Case 2 in the pre-pubertal period was obese (BMI Z score +2.1) with small testes volumes (1.5 mL bilaterally) and micropenis. He had appropriately low basal gonadotropins and serum testosterone for age, with normal inhibin B and low AMH. Abbreviations: Tvol, testes volume; G stage, Tanner genital stage.

|  | Age<br>(yrs) | Tvol<br>(mL) | G<br>stage | BMI | Basal LH<br>(IU/L)<br>(0.1-0.6) | Basal<br>FSH<br>(IU/L)<br>(0.1-0.9) | Peak<br>LH | Peak<br>FSH | Testosterone<br>(nmol/L) | Inhibin B<br>(pg/mL)<br>(55-255) | AMH<br>(ng/mL) | TSH<br>(mIU/L)<br>(0.3-4) | T4<br>(pmol/L) |
| --- | --- | --- | --- | --- | --- | --- | --- | --- | --- | --- | --- | --- | --- |
| **Case 1** | **Clinical data at first assessment** |  |  |  |  |  |  |  |  |  |  |  |  |
|  | 17-21.9 | 10 | 3 | 36 | 1.5 | 3.1 | 16.6 | 6.6 | 0.7 | 142.3 | - | 4.5 | 15.2 |
|  | **Clinical data after therapy with i.m. testosterone esters** |  |  |  |  |  |  |  |  |  |  |  |  |
|  | 22-26.9 | 12 | 4 | 38 | <0.3 | 0.2 | - | - | 7.4 | - | - | 6 | 15.5 |
| **Case 2** | **Clinical data at first assessment** |  |  |  |  |  |  |  |  |  |  |  |  |
|  | 6-11 | 1.5 | 1 | 22 | 0.6 | 1.5 | - | - | 0.8 | 213.1 | 49.3 | 2.5 | - |

Case 2 (NM_181303.1: c.163C>T; p.R55*) is a pre-pubertal boy who presented with stereotypia, learning difficulties, ASD features and micropenis, which is a typical predictive sign of HH in male infants (3). At the age of 7-12 years, he was obese (BMI Z-score +2.1, height 148 cm) with small testes volumes (1.5 mL bilaterally). He was also found to have corpus callosum dysplasia (data not shown). His parent also had history of learning difficulties and stereotypic behaviors, but there was no family history of GD. He had appropriately low gonadotropins and testosterone for age, with normal inhibin B and low AMH (**Table 2**).

In both cases, exome sequencing of the probands excluded deleterious variants in known GD genes and Sanger sequencing of their parents confirmed an heterozygous carrier in each family. DNA from other Case 2 family members was not available.

Interestingly, both variants cause premature stop codons within the *NLGN3* extracellular domain, resulting in truncated proteins which are likely to be dysfunctional. Both variants are novel, not found in the gnomAD database (v2.1.1, accessed 06.02.2022) and, notably, the *NLGN3* gene is highly intolerant to protein-truncating changes (pLi 0.98). According to American College of Medical Genetics (ACMG) (23), these variants are both classified as pathogenic (**Figure 5B**). Bioinformatic tools predicted these variants to be damaging, deleterious and disease causing, with CADD score of >35 and DANN score of >0.99 (**Figure 5C and Table 3**). Further, multi-species protein alignment combined with GERP++ software showed that identified variants affect highly conserved residues (**Figure 5C**).

**Table 3.** Bioinformatic tools applied to predict pathogenicity of *NLGN3* variants. Chromosome position, nucleotide substitution, amino acid substitution and bioinformatic predictions of the identified *NLGN3* variants.

|  | Chr | Position<br>GRCh37/hg19 | nt sub<br>NM_181303.1 | aa sub<br>NP_851820.1 | FATHMM-MKL | LRT | Mutation<br>Taster | DANN | ACMG<br>verdict |
| --- | --- | --- | --- | --- | --- | --- | --- | --- | --- |
| **Case 1** | X | 70367965 | c.366 G>A | p.W122* | Damaging | Deleterious | Disease<br>causing | 0.9968 | Pathogenic |
| **Case 2** | X | 70367762 | c.163 C>T | p.R55* | Damaging | Deleterious | Disease<br>causing | 0.9981 | Pathogenic |

### NLGN3 is upregulated in maturing GnRH neurons and NLGN3 truncating variants impair protein synthesis and neurite outgrowth in vitro

Our transcriptomic analyses revealed that the X-linked *NLGN3* gene was up-regulated in primary and immortalized GnRH neurons during development (**Figure 4B**). Thus, to validate its developmentally regulated expression, we performed the following experiments. First, we confirmed higher *NLGN3* levels in maturing GT1-7 versus immature GN11 cells (**Figure 6A,B**), by qRT-PCR and immunocytochemistry, using a previously validated anti-NLGN3 antibody. Then, we analyzed NLGN3 expression on E14.5 mouse sections and confirmed co-localisation of NLGN3 protein in GnRH neurons that had settled in the maturing hypothalamus (**Figure 6C**).

**Figure 6.**
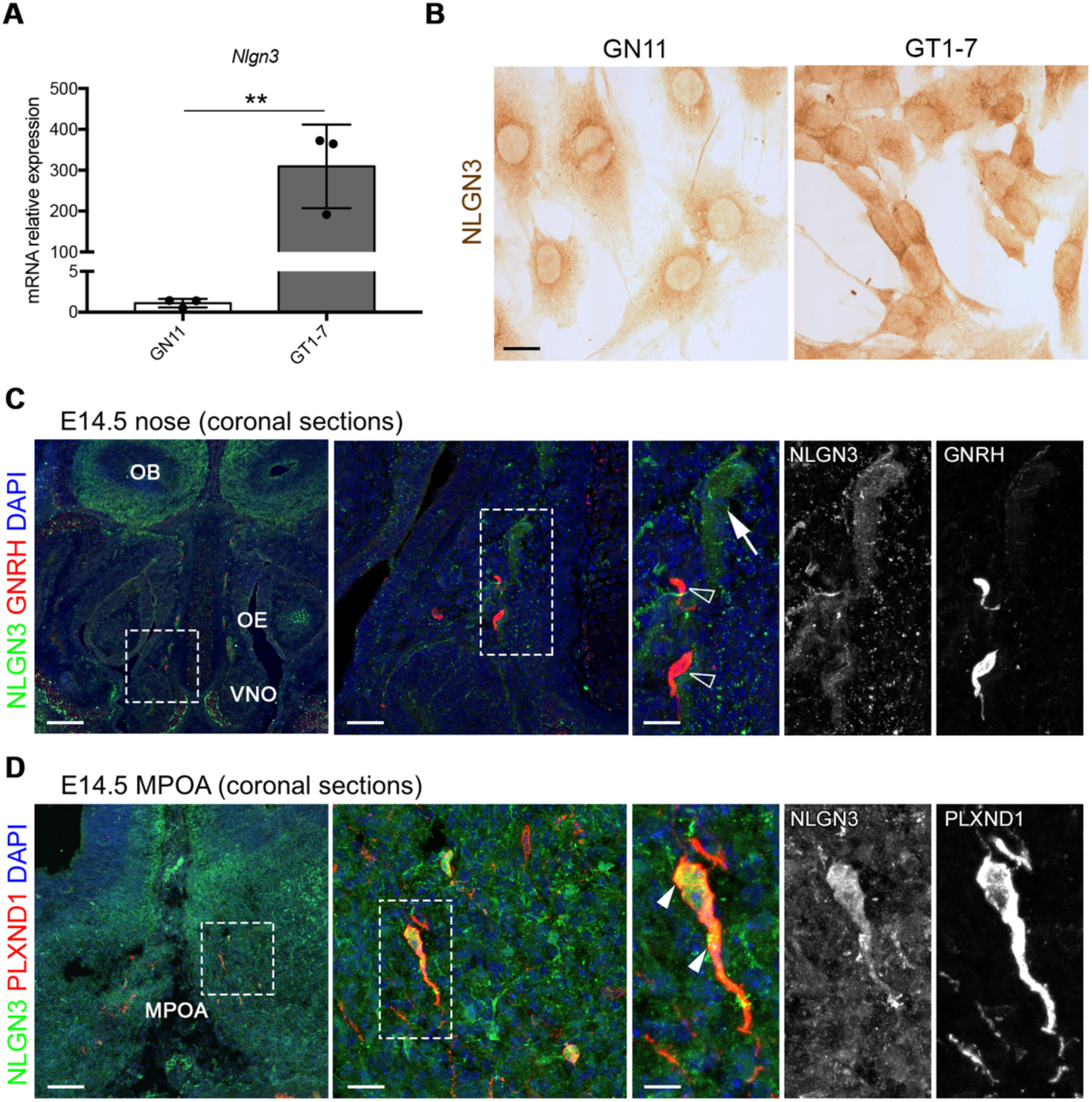
NLGN3 is developmentally regulated in GnRH neurons. A - qPCR analysis performed on GN11 and GT1-7 cells revealed higher *Nlgn3* expression levels in GT1-7 cells (log_FC_ = 8.12, P < 0.01). This result is in line with microarray experiments (log_FC_ = 4.15; P < 0.00001). Data are presented as mean ± SD of 3 biological replicates. P values indicate Student’s t test (** P < 0.01). B - Immunoperoxidase staining for NLGN3 on GN11 and GT1- 7 revealed different levels of endogenous NLGN3 protein in these cells. Scale bar: 25 μm. C,D - Coronal sections of E14.5 mouse heads were immunolabelled for NLGN3 together with GnRH or PLXND1 to detect GnRH neurons. Sections are shown at the level of the VNO (nose, C) or MPOA (forebrain, D). White boxes indicate areas shown at higher magnification on the right of the corresponding panel, with single channels shown also adjacent to the panel. Open arrowheads indicate examples of GnRH-positive cells that lack NLGN3. Arrows indicate examples of NLGN3-positive cells in the nasal parenchyma (C). Arrowheads indicate examples of GnRH-positive cells with NLGN3 (D). Sections were counterstained with DAPI. OE, olfactory epithelium; OB, olfactory bulb; VNO, vomeronasal organ; MPOA, medial preoptic area. Scale bars: 250 (right panels), 150 (middle panels) or 50 μm (left panels).

Then, to functionally validate the predicted pathogenicity of the identified *NLGN3* variants, we overexpressed human HA-tagged wild type (WT) (24), R55* or W122* NLGN3 in cell culture models. Initially, protein synthesis and secretion of WT versus mutated NLGN3 proteins were evaluated in COS7 cells, by immunoblotting analysis on lysates and conditioned media, respectively. As expected, a 110 kDa band corresponding to the full-length protein was observed in lysates of COS7 cells transfected with NLGN3 WT (**Figure 7A**) (25). In contrast, R55* and W122* variants led to the formation of prematurely truncated proteins of expected molecular weights (7 and 14 kDa, respectively) (**Figure 7A**). In addition, a shorter form (90 kDa) of NLGN3 WT, which likely represents the ectodomain released by proteolytic shedding (25), was detected in conditioned media. We could not detect equivalent shed or secreted forms for the NLGN3 mutant proteins (**Figure 7A**). To study localization of these mutant proteins, we performed similar overexpression experiments and immunocytochemical analysis in GN11 cells, which show low levels of endogenous NLGN3 (**Figure 6A,B**). Immunofluorescence staining for HA-tag in GN11 cells revealed that mutant proteins were only detected in the endoplasmic reticulum (ER), selectively labeled with mEmerald-ER-3 vector (**Figure 7B**). By comparison, the WT protein was efficiently transported to the plasma membrane. Of note, we observed only very few HA^+^ cells when transfected with R55* NLGN3, strongly supporting a rapid NLGN3 protein degradation and consequent lack of detection, as previously described for other NLGN3 mutant proteins (26). Altogether, these observations strongly support a LoF effect of the two newly identified variants.

**Figure 7.**
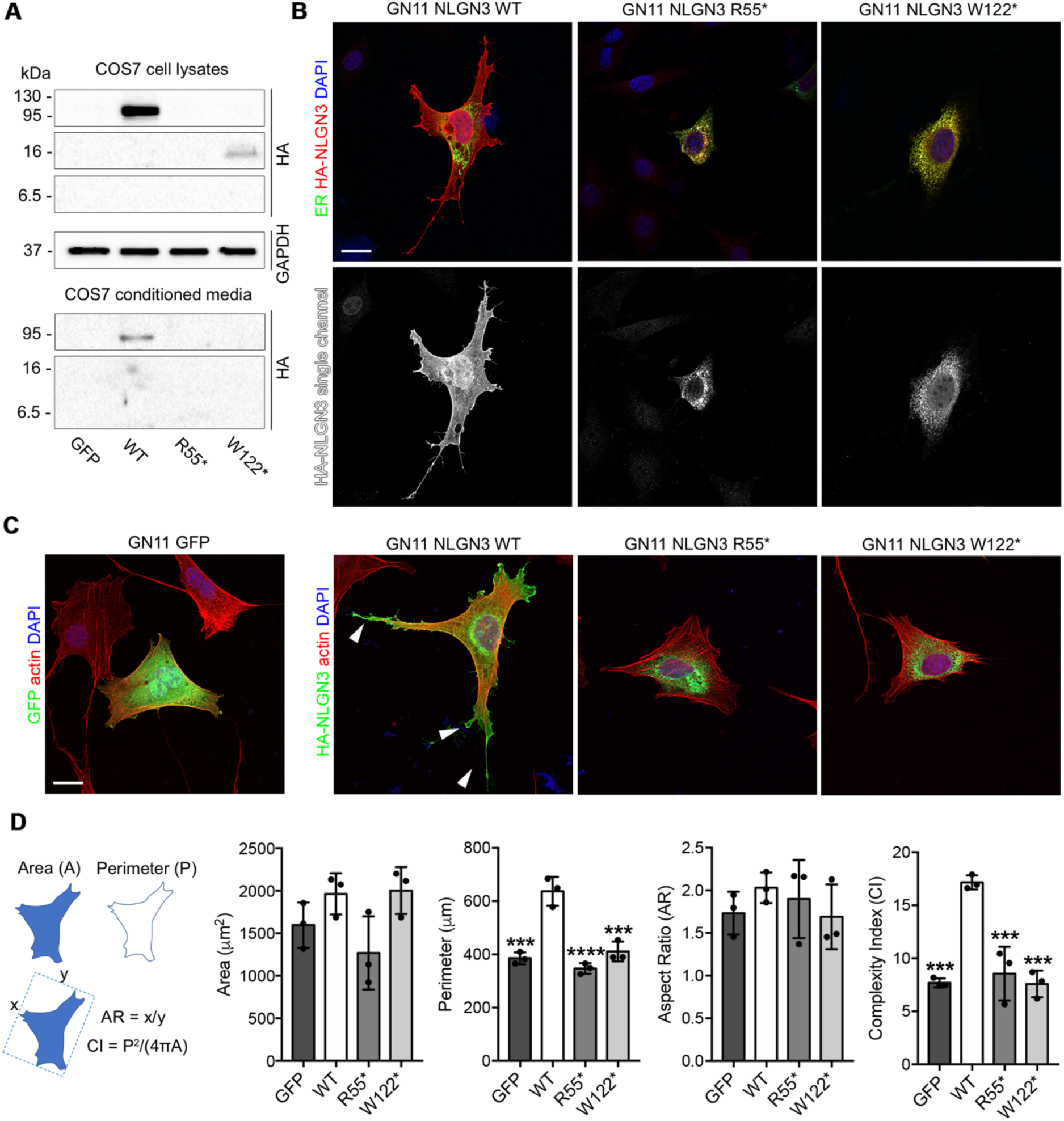
Mutant NLGN3 proteins induce ER-retention and impair neuritogenesis in immortalized GnRH neurons. A - Immunoblot analysis with anti-HA or anti-GAPDH antibodies on whole-cell lysates and conditioned media from COS7 cells overexpressing control vector (GFP), HA-tagged WT or mutant NLGN3. GAPDH is shown as a loading control for cell lysates. B - Confocal images of GN11 cells transfected cells with mEmerald-ER-3 (green) and human WT or mutated HA-tagged NLGN3 vectors and stained for HA (red) after 24 h (right panels). HA-NLGN3 single channel images are shown below each panel. C - Confocal images of GN11 transfected with human WT or mutated NLGN3 HA-tagged encoding vector and stained for HA (green) and F-actin (red). Arrowheads point at neurites in NLGN3 WT-expressing GN11 cells. D - Quantification of cell perimeter (P), cell area (A), aspect ratio (AR) and complexity index (CI) in GN11 cells transfected with indicated plasmids. Column graph quantification showing a significant increase in the number of neurites extending from NLGN3 WT-expressing cells compared to NLGN3 mutants and GFP-transfected control cells. Data are presented as mean ± SD of 3 biological replicates. Significant differences were determined by one-way ANOVA followed by Tukey’s multiple comparison test (***P < 0.001; ****P<0.0001). Nuclei were counterstained with DAPI. Scale bar: 25 μm.

Because NLGN3 promotes neuritogenesis in human neural progenitor cells (27), we studied the effect of WT or mutant NLGN3 proteins on GnRH neuron morphology and neurite outgrowth by overexpression experiments in GN11 cells, as previously described (28). NLGN3 WT promoted the formation of several protrusions, with cells displaying an increased spreading compared to GFP-transfected controls (**Figure 7C**). By contrast, the two mutants (R55* and W122*) were unable to induce such changes (**Figure 7C**), with transfected cells displaying a significantly lower cell perimeter, although total area and cell polarity (i.e. aspect ratio) were not affected (**Figure 7D**). Further, cells overexpressing NLGN3 mutants exhibited a significantly reduced cell shape complexity (i.e. complexity index) compared to NLGN3 WT, revealing a strong defect in cell protrusion formation (**Figure 7D**). Together, these results confirmed that the identified NLGN3 variants are LoF and failed to induce cytoskeletal and membrane rearrangements typical of neuritogenesis.

## Discussion

In this work, we have revealed for the first time the transcriptome profiles of developing primary GnRH and immortalized neurons and provide a new resource for understanding the key developmental regulation of GnRH neurons. Further, we show the role of NLGN3 in the development of GnRH neurons and identify novel *NLGN3* LoF variants in two unrelated patients with GD and ASD traits.

Interestingly, our transcriptomic data highlight the presence of genetic signatures similar to human iPSCs-derived GnRH neurons (9), strongly supporting the high conservation of human and rodent GnRH neuron systems.. Further, the presence among DEGs of genes experimentally associated with GnRH neuron development (e.g. *Reln*, *Cxcr7*) (14) or with GD pathogenesis (e.g. *Sema3a*, *Fgfr1*) (4) emphasizes the efficacy of dissecting the genetic signatures of GnRH neurons in rodents to decipher the molecular mechanisms of GnRH neuron biology and to predict human disease genes.

As an exemplar, our integrated *omic* analysis allowed us to identify *NLGN3* as a new candidate GD gene. Specifically, the identified variant in our Case 1 proband induces a premature stop codon in *NLGN3*, a gene that has been linked so far to ASD (12, 26, 29, 30) but not to GD. Notably, the other patient identified in this study (Case 2) is also hemizygous for a nonsense *NLGN3* variant and, although prepubertal, displayed typical red flag signs of hypogonadism, including small testes volume and micropenis (3). In addition, these two probands had behavioral difficulties including ASD features or NDD, supporting the existence of a shared pathophysiological link between GD and other developmental disorders, as hypothesized by a recent phenotypic population study showing an increased risk of NDDs in patients with GD (31). This implies that NLGN3 may be essential for the development or function of GnRH neurons, besides its well-known role in brain neurotransmission, which is thought to be dysfunctional in ASD (12). Although its expression has been reported in the developing telencephalon of chick and zebrafish embryos in territories relevant to GnRH neuron development (32, 33), the role of NLGN3 in this system is not yet known.

In this context, because NLGN3 is developmentally up-regulated both in primary and immortalized GnRH neurons and its overexpression in GN11 cells promotes neuritogenesis, NLGN3 may be required to promote neurite extension during the final phases of GnRH neuron development *in vivo*. Consequently, NLGN3 loss might have an impact on this process, thus causing GD. Interestingly, compound double *Nlgn1/3-* or *Nlgn2/3*-null mice display a reduced reproductive rate (34), in addition to behavioral phenotypes related to ASD (35), strongly supporting a role of NLGN3 in the reproductive axis. Yet, future detailed phenotypic analyses of GnRH neuron development in NLGNs single and compound null mice will be necessary to define the exact function of NLGN3 in these systems.

In addition, our study also links for the first time nonsense *NLGN3* variants to a combined phenotype of ASD and GD. Most ASD patients with mutated *NLGN3* carry missense variants, with only two previous reports of nonsense variants without functional characterization or phenotypic correlation (12). Further, although we cannot exclude that the identified nonsense *NLGN3* variants undergo degradation *in vivo* due to the nonsense-mediated decay mechanism of RNA surveillance, our data still provide evidence that if this mechanism does not occur, remaining NLGN3 mutant proteins are non-functional, and their expression leads to a relevant cellular phenotype.

Although the association of truncating *NLGN3* variants with the co-existence of GD with ASD or NDD is being supported so far only by the two cases reported in our study, it will be interesting to expand genotype-phenotype correlation to other cases and to understand the molecular mechanisms underlying this association. Interestingly, growing evidence supports the existence of common genetic determinants between NDDs and GD, as shown by a recent study where *SOX11* mutations have been linked to a novel NDD with HH (36).

Overall, our study, by exploiting the power of *omic* analyses, has provided a novel experimental resource which will be invaluable for the identification of the elusive mechanisms underlying GnRH neuron biology and related diseases. Further, our findings provide the first proof-of-principle that the transcriptomic analysis of primary and immortalized GnRH neurons, combined with human genetics, *in silico* tools and *in vitro* models, may be applied to identify novel GD candidate genes. In the future, mutational screening of our top 20 genes in other patient cohorts and further analysis of GnRH neuron transcriptomes might be exploited to identify novel genetic determinants of GD. Last, because our studies imply the existence of a genetic pathway shared between ASD and GD, it will be important for clinicians to be aware of the potential for pubertal and reproductive disorders in children with ASD/NDD to identify those who might require hormonal therapy intervention and, vice versa, to assess the psychological and social aspects of GD-affected children reporting ASD or developmental delay.

## Materials and methods

### Animals

*Gnrh1*-GFP rat embryos (7) were used in Wistar background to isolate GnRH-GFP neurons. Wild type C57/Bl6 mouse embryos were used for expression studies (Italian Ministry of Health, license N° 5247B.N.QPE). To obtain embryos of defined gestational stages, rats and mice were mated in the evening and the morning of vaginal plug formation was counted as E0.5.

### Cell lines

GN11, GT1-7 cells (5) and COS7 cells were grown as a monolayer at 37°C in a humidified CO_2_ incubator in complete DMEM (Euroclone) supplemented with 10% fetal bovine serum (FBS; Invitrogen). Subconfluent cells were harvested by trypsinization and cultured in 57 cm^2^ dishes. Cells within six passages were used for all experiments.

### Cell dissociation and fluorescence-activated cell sorter (FACS) analysis

Explants from the nasal area (E14) and from nasal and forebrain areas (E17-20) were microdissected from heads of *Gnrh1*-GFP embryos; nose and forebrain cells were dissociated by incubation in 0.05% trypsin with 100 ug/mL DNase I in neurobasal medium (Invitrogen) at 37°C for 15 minutes. Trypsinization was quenched by addition of neurobasal medium containing 10% heat-inactivated fetal bovine serum (FBS; Invitrogen) at 37°C for 5 minutes. Cells were washed 3 times in neurobasal medium (without FBS) to remove serum before FACS and resuspended in neurobasal medium without phenol red (Invitrogen) containing L-glutamine (Invitrogen) and B-27 supplement (1:50; Invitrogen). Dissociated cells from 8 - 10 embryos for each age were pooled for each FACS. FACS was performed by the Wolfson Scientific Support Services (UCL, London, UK) by using a MoFlo Sorter (Dako). A non-green embryo was used as a control for fluorescence. Cells were excited by using a 488-nm Argon laser and detected by using a 530/40 (FL1) bandpass filter. A cell purity of 95–98.5% was obtained for each sort. Sorted (GFP^+^) and unsorted (GFP^-^) cells were directly collected in lysis buffer (Qiagen) and used for RNA extraction.

### Microarray analysis

For GN11 and GT1-7 cells, total RNA was isolated by acid guanidinium thiocyanate-phenol-chloroform extraction (Trizol, Invitrogen) followed by a Qiagen RNeasy kit clean-up procedure (Qiagen). Total RNA from GFP^+^ and GFP^-^ FACS-purified cells was extracted immediately after collection by using the Qiagen RNeasy Plus kit (Qiagen). RNA integrity was verified with RNA 6000 Nano and Pico kit respectively and only high quality RNAs, with RIN greater than 7, were used. WT Expression Kit (Invitrogen) and Ovation Pico WTA System V2 (Nugen) were respectively used to prepare sense-strand cDNA. Biotin labeling was performed with either WT Terminal Labeling kit Affymetrix (Invitrogen) and Encore Biotin Module kit (Nugen). Labeled cDNA was hybridized (45°C for 17 hours) to the Affymetrix Mouse Gene 1.0 ST and Rat Gene 1.0 ST Gene chips according to manufacturer’s protocol.

### Data processing and visualization

Microarray data analysis, including quality controls, normalization and gene filtering was conducted using AMDA software (37). The identification of DEGs was addressed using linear modeling approach and empirical Bayes methods (38) together with FDR correction of the *P* value (Benjamini-Hochberg); the adjusted *<u>P</u>* value that has been selected in the analysis is <0.05 and log_FC_ > 2. Gene ontology analyses were performed as previously described with reSTRING software (15). Briefly, hierarchical clustering was performed with Euclidean metric on log-transformed ratios of probes fluorescence intensity to average probe intensity (39). PCA was performed with Scikit-learn (40). Data visualization was performed with SciPy (39), matplotlib (41) and seaborn (42) libraries for the Python programming language.

### Human samples and sequencing

Case 1. Exome sequencing data from 47 patients from a UK GD cohort (CPMS ID 30730) were analyzed. Exome sequencing was performed on DNA extracted from peripheral blood leukocytes, using an Agilent V5 platform and Illumina HiSeq 2000 sequencing. The exome sequences were aligned to the UCSC hg19 reference genome using the Burrows-Wheeler Aligner software (BWA-MEM [bwa-0.7.12]. Picard tools [picard- tools-1.119] was used to sort alignments and mark PCR duplicates. The genome analysis toolkit (GATK-3.4-46) was used to realign around indels and recalibrate quality scores using dbSNP, Mills and 1,000 genomes as reference resources. Variant calling and joint genotyping using pedigree information was performed using HaplotypeCaller in GVCF mode from the genome analysis toolkit. The resulting variants were filtered using the variant quality score recalibration (VQSR) function from GATK. An analysis of the called variants was performed using Ingenuity Variant Analysis (QIAGEN Redwood City, www.qiagen.com/ingenuity). Filtering for potential causal variants was carried out using filters for quality control, allele frequency and predicted functional annotation. Potentially pathogenic variants in candidate genes were verified by Sanger sequencing. Libraries of genomic DNA samples were prepared using the Agilent Sureselect Human All Exon v5 kit (Agilent Technologies), and were sequenced on a HiSeq instrument (Illumina) according to the manufacturer’s recommendations for paired-end 76-bp reads. The bioinformatics pipeline, alignment processes, and quality procedures were as previously described (43). Version 3.4–46 of the Genome Analysis Toolkit was used for this study.

Case 2. Exome sequencing was realized as previously published (44). Briefly, libraries of genomic DNA samples were prepared using the Twist Human Core Exome kit (Twist Biosciences), and were sequenced on a NovaSeq 6000 instrument (Illumina) according to the manufacturer’s recommendations for paired-end 151-bp reads. A mean depth of 86.96 x was reach and 97.2 % of the refseq exons were covered at least by 10 reads. Variants were identified using a computational platform of the FHU Translad, hosted by the University of Burgundy Computing Cluster (CCuB). Raw data quality was evaluated by FastQC software (v0.11.4). Reads were aligned to the GRCh37/hg19 human genome reference sequence using the Burrows-Wheeler Aligner (v0.7.15) (45). Aligned read data underwent the following steps: (a) duplicate paired-end reads were removed by Picard software (v2.4.1), and (b) base quality score recalibration was done by the Genome Analysis Toolkit (GATK v3.8) Base recalibrator. Using GATK Haplotype Caller, Single Nucleotide Variants with a quality score >30 and an alignment quality score >20 were annotated with SNPEff (v4.3) (46). Rare variants were identified by focusing on nonsynonymous changes present at a frequency less than 1% in the GnomAD database. Copy Number Variants were detected using xHMM (v1.0), were annotated using in-house python scripts and then filtered regarding their frequency in public databases (DGV, ISCA, DDD).

### Expression vectors and transfection

To introduce the c.163C>T and c.366G>A variants into human *NLGN3* gene, the WT human HA-tagged NLGN3 expression vector (Addgene, 59318) was mutagenized using the QuickChange Lightning Site-Directed Mutagenesis Kit (Agilent Technologies) and specific oligonucleotides for *NLGN3* R55* (FW 5’- GGCAGTGGTACTCAGGCACCCCTTAGC-3’, RV 5’- GCTAAGGGGTGCCTGAGTACCACTGCC-3’) and W122* (FW 5’- GTCATGCTGCCGGTCTGATTCACTGCCAACTTGGATATCG-3’, RV 5’- TCAGACCGGCAGCATGACTTCGGGCACAGCTGTGTGGATG-3’) respectively. To visualize the ER, the mEmerald-ER-3 vector (gift from Prof. Diego De Stefani; Addgene, 54082) was co-transfected. Expression vectors were transiently transfected (for 24 hours) into GN11 cells using Lipofectamine 3000 (Invitrogen).

### Immunoblotting

Cells were lysed in 150 mM NaCl, 50 mM Tris-HCl (pH 7.4), and 1% Triton X-100, supplemented with protease and phosphatase inhibitors (Roche). 20 μg of proteins were transferred to nitrocellulose membranes (Bio-Rad), immunoblotted with rabbit anti–HA (1:1,000; Cell Signaling, 3724) and rabbit anti-GAPDH (1:5,000; Cell Signaling, 2118), followed by HRP-conjugated anti-rabbit antibody (1:10,000; Santa Cruz Biotechnology Inc., sc-2301).

### Immunocytochemistry

Paraformaldehyde-fixed GN11 cells were incubated with PBS containing 10% normal goat serum and 0.1% Triton X-100 Rabbit anti-HA (1:800; Cell Signaling, 3724) and chicken anti-GFP (1:1000; Abcam, ab13970) were used as primary antibodies. Secondary antibodies used were 488-conjugated donkey anti-rabbit and anti-chicken and Cy3-conjugated donkey anti-rabbit Fab fragments (1:200; Jackson Immunoresearch). To detect F-actin, cells were stained with TRITC-conjugated phalloidin (1:400; Sigma-Aldrich, P1951) for 30 min at 37 °C(47). Nuclei were counterstained with DAPI (1:10,000; Sigma-Aldrich, D9542). For immunoperoxidase labeling, cells were incubated with hydrogen peroxide to quench endogenous peroxidase activity before incubation with rabbit anti-NLGN3 (1:200), followed by biotinylated goat anti-rabbit antibody (1:400; Vector Laboratories, BA-1000), and then developed with the ABC kit (Vector Laboratories, PK6100) and DAB (Sigma-Aldrich, D4293).

### RT-PCR and qPCR

Total RNA from GN11, GT1-7 and FACS-sorted cells was retrotranscribed into cDNA as previously described (48). Briefly, 1 μg of total RNA was reverse transcribed with random hexamers and MultiScribe reverse transcriptase (Applied Biosystems) by following the manufacturer’s instructions. The expression level of murine *Nlgn3 (*fw 5’-GAAGATGGATCCGGCGCTAA-3’; rev 5’-ACGATGACGTTGCCGTAACT- 3’*), Pdgfra (*fw 5’-CGCTGGAGGGTTATCGAGTC-3’; rev 5’- CAGGTTGGGACCGGCTTAAT-3’*), L1cam (*fw 5’-GCTCCTCATCCTGCTCATCC-3’; rev 5’-TCTCCAGGGACCTGTACTCG-3’*), Sema3c (*fw 5’-GAACCCATGTTTGTGGACGC-3’; rev 5’-CCACCAGTGTCATTAGGGCA-3’) and *Tgfb1* (fw 5’- CGCAACAACGCCATCTATGA-3’; rev 5’*-*ACTGCTTCCCGAATGTCTGA-3’) *genes* was quantified by qPCR on a Biorad CFX Connect thermal cycler with Luna Universal qPCR Master Mix (NEB) in 10 μL reactions, with final concentration of 0.25 μM for each primer. The cycling conditions were 95 °C for 1 min, followed by 40 cycles of 15 s at 95 °C, 30 s at 60 °C and 30 s at 72 °C. A final melting curve analysis assured the authenticity of the target product. Triplicate samples were run in all reactions; first-strand DNA synthesis reactions without reverse transcriptase were used as controls. The ΔCq value and the ΔΔCq were calculated relative to control samples using quantification cycle (Cq) threshold values that were normalized to the reference gene, *Gapdh* (fw: 5′-CATCCCAGAGCTGAACG-3′; rev 5′- CTGGTCCTCAGTGTAGCC-3′).

### Immunostaining

Wild type C57/Bl6 embryos at E14.5 were fixed for 3 hours in 4% paraformaldehyde (PFA) and then cryoprotected overnight in 30% sucrose, embedded in OCT and cryosectioned for immunostaining. PFA-fixed tissue sections (25 μm) or cells were incubated with with serum-free protein block (DAKO, X0909). For immunofluorescence staining, goat anti-NLGN3 (1:25; Santa-Cruz, sc-14091), rabbit anti-GnRH (1:400, Immunostar, 20075), rabbit anti-NLGN3 (1:100) (49), goat anti-PLXND1 (1:200; R&D, AF4160) (50) were used as primary antibodies. Secondary antibodies used were 488- conjugated donkey anti-rabbit and Cy3-conjugated donkey anti-goat Fab fragments (1:200; Jackson Immunoresearch). Nuclei were counterstained with DAPI (1:10,000; Sigma-Aldrich, D9542).

### Image acquisition

Cells were examined with a Zeiss LSM 900 confocal laser scanning microscope equipped with a Zeiss Axiocam 305 color and using a Plan Apochromat 40X 1.3 Pil DIC (VIS-IR M27) oil immersion objective (Zeiss). DAPI, Alexa488 and Cy3 were excited at 353, 493 and 548 nm and observed at 400-500, 500-540 and 540- 700 nm, respectively. 1024x1024 pixels images were captured in a stepwise fashion over a defined z-focus range corresponding to all visible fluorescence within the cell. Maximum projections of the z-stack with 0.25 μm optical section were obtained post-acquisition by using Zeiss ZEN System. Adobe Photoshop CS6 software was used to prepare the presented images.

### Quantification

Morphological analysis were performed on confocal images using ImageJ (v1.52a) as previously described (28) and summarized in Fig. 4D.

### Statistics

Statistical tests employed are outlined in figure legends and were conducted when the experiment had been performed a minimum of 3 times on a minimum of 3 individual samples. Data are presented as mean ± SD and results considered significant with a *P* value less than 0.05 (GraphPad Prism 7.0).

### Study approval

Human and animal approval: Ethical approval for Case 1 was granted by the London-Chelsea NRES committee (13/LO/0257) and the UK NHS Health Research Authority (IRAS 95781). Ethical approval for Case 2 was granted by Ethics Committees of CHU de Caen and CHU Dijon Bourgogne. All participants provided written informed consent prior to study participation. The study was conducted in accordance with the guidelines of The Declaration of Helsinki. All individual-level data, including clinical data, was de-identified. Participants or their legal representatives gave consent to the publication of the results of this research work in the present study. The animal work was approved by the University of Milan Animal Welfare Body and by the Italian Minister of Health to AC and was conducted in accordance with the EU Directive 2010/63/EU.

## Data Availability

The exome sequencing datasets generated during and/or analyzed during the current study are available from the corresponding author on reasonable request.

## Acknowledgements

We would like to thank: Professor Maria Foti and Genopolis gene-sequencing facility (University of Milano-Bicocca) for their help with Affymetrix microarrays and analyses; Dr. Thomas Adejumo for help with FACS at the UCL Wolfson Institute for Biomedical Research; Professor John Parnavelas for lab resources. We also thank the imaging facility NOLIMITS at the University of Milan for help with confocal imaging.

## Author contributions

Conceptualization: A.C., S.R.H., V.A.

Data curation: R.O., A.L., S.M., M.B., S.R.H

Formal analysis: R.O., A.L., S.M., P.D.

Funding acquisition: A.C., S.R.H., R.O., V.V.

Investigation: R.O., A.L., A.P., P.G., P.D., F.A., H.L.S., V.B., C.P., A.V.

Resources: F.M., H.L.S., P.S.

Software: S.M., M.B.

Supervision: A.C., S.R.H., V.M., V.V.

Visualization: R.O., S.M.

Writing – original draft: R.O., A.L., A.C., S.H.

## Web resources

RESTRING: https://github.com/Stemanz/restring

ToppGene: https://toppgene.cchmc.org/ Gene Matcher: https://genematcher.org/ Varsome: https://varsome.com/

## Data availability

Microarray data will be publicly released upon manuscript acceptance on GEO NCBI. Access tokens will be released to reviewers if requested

Potentially identifying clinical information (for example age at assessment and investigation) has been removed to comply with medRxiv publication requirements but is available from the corresponding author.

## References

1. Herbison AE. Control of puberty onset and fertility by gonadotropin-releasing hormone neurons. Nat. Rev. Endocrinol. 2016;12(8):452–466.

2. Oleari R, et al. PLXNA1 and PLXNA3 cooperate to pattern the nasal axons that guide gonadotropin-releasing hormone neurons. Development 2019;146(21). doi:10.1242/dev.176461

3. Boehm U, et al. Expert consensus document: European Consensus Statement on congenital hypogonadotropic hypogonadism--pathogenesis, diagnosis and treatment.. Nat. Rev. Endocrinol. 2015;11(9):547–64.

4. Oleari R, et al. The Differential Roles for Neurodevelopmental and Neuroendocrine Genes in Shaping GnRH Neuron Physiology and Deficiency. Int. J. Mol. Sci. 2021;22(17):9425.

5. Maggi R, et al. Immortalized luteinizing hormone-releasing hormone neurons show a different migratory activity in vitro.. Endocrinology 2000;141(6):2105–12.

6. Messina A, et al. A microRNA switch regulates the rise in hypothalamic GnRH production before puberty.. Nat. Neurosci. 2016;19(6):835–44.

7. Kato M, et al. Characterization of voltage-gated calcium currents in gonadotropin-releasing hormone neurons tagged with green fluorescent protein in rats.. Endocrinology 2003;144(11):5118–25.

8. Abraham E, et al. The zebrafish as a model system for forebrain GnRH neuronal development. Gen. Comp. Endocrinol. 2009;164(2–3):151–160.

9. Lund C, et al. Characterization of the human GnRH neuron developmental transcriptome using a GNRH1-TdTomato reporter line in human pluripotent stem cells.. Dis. Model. Mech. 2020;13(3):dmm040105.

10. Uchigashima M, Cheung A, Futai K. Neuroligin-3: A Circuit-Specific Synapse Organizer That Shapes Normal Function and Autism Spectrum Disorder-Associated Dysfunction. Front. Mol. Neurosci. 2021;14(October):1–22.

11. Xu J, et al. Neuroligin 3 Regulates Dendritic Outgrowth by Modulating Akt/mTOR Signaling.. Front. Cell. Neurosci. 2019;13(November):518.

12. Nguyen TA, Lehr AW, Roche KW. Neuroligins and Neurodevelopmental Disorders: X-Linked Genetics.. Front. Synaptic Neurosci. 2020;12(August):33.

13. Cariboni A, Maggi R, Parnavelas JG. From nose to fertility: the long migratory journey of gonadotropin-releasing hormone neurons. Trends Neurosci. 2007;30(12):638–644.

14. Cho H-J, et al. Nasal Placode Development, GnRH Neuronal Migration and Kallmann Syndrome. Front. Cell Dev. Biol. 2019;7(July):1–27.

15. Manzini S, et al. reString: an open-source Python software to perform automatic functional enrichment retrieval, results aggregation and data visualization.. Sci. Rep. 2021;11(1):23458.

16. Forni PE, Wray S. GnRH, anosmia and hypogonadotropic hypogonadism--where are we?. Front. Neuroendocrinol. 2015;36(2):165–77.

17. Van Battum EY, Brignani S, Pasterkamp RJ. Axon guidance proteins in neurological disorders. Lancet Neurol. 2015;14(5):532–546.

18. Chen J, et al. ToppGene Suite for gene list enrichment analysis and candidate gene prioritization.. Nucleic Acids Res. 2009;37(Web Server issue):W305-11.

19. Maione L, et al. GENETICS IN ENDOCRINOLOGY: Genetic counseling for congenital hypogonadotropic hypogonadism and Kallmann syndrome: new challenges in the era of oligogenism and next-generation sequencing.. Eur. J. Endocrinol. 2018;178(3):R55–R80.

20. Saengkaew T, et al. Genetic evaluation supports differential diagnosis in adolescent patients with delayed puberty. Eur. J. Endocrinol. 2021;185(5):617–627.

21. Ferreira L, Silveira G, Latronico AC. Approach to the patient with hypogonadotropic hypogonadism. J. Clin. Endocrinol. Metab. 2013;98(5):1781–1788.

22. Sobreira N, et al. GeneMatcher: A Matching Tool for Connecting Investigators with an Interest in the Same Gene. Hum. Mutat. 2015;36(10):928–930.

23. Richards S, et al. Standards and guidelines for the interpretation of sequence variants: a joint consensus recommendation of the American College of Medical Genetics and Genomics and the Association for Molecular Pathology. Genet. Med. 2015;17(5):405– 423.

24. Chih B, et al. Disorder-associated mutations lead to functional inactivation of neuroligins.. Hum. Mol. Genet. 2004;13(14):1471–7.

25. Bemben MA, et al. Isoform-specific cleavage of neuroligin-3 reduces synapse strength. Mol. Psychiatry 2019;24(1):145–160.

26. Quartier A, et al. Novel mutations in NLGN3 causing autism spectrum disorder and cognitive impairment. 2019:

27. Gatford NJF, et al. Nanoscopic Clustering of Neuroligin-3 and Neuroligin-4X Regulates Growth Cone Organization and Size. bioRxiv [published online ahead of print: 2019]; doi:10.1101/546499

28. Bouilly J, et al. DCC/NTN1 complex mutations in patients with congenital hypogonadotropic hypogonadism impair GnRH neuron development.. Hum. Mol. Genet. 2018;27(2):359–372.

29. Jamain S, et al. Mutations of the X-linked genes encoding neuroligins NLGN3 and NLGN4 are associated with autism. Nat. Genet. 2003;34(1):27–29.

30. Redin C, et al. Efficient strategy for the molecular diagnosis of intellectual disability using targeted high-throughput sequencing. J. Med. Genet. 2014;51(11):724–736.

31. Ohlsson Gotby V, et al. Hypogonadotrophic hypogonadism, delayed puberty and risk for neurodevelopmental disorders. J. Neuroendocrinol. 2019;31(11). doi:10.1111/jne.12803

32. Davey C, Tallafuss A, Washbourne P. Differential expression of neuroligin genes in the nervous system of zebrafish.. Dev. Dyn. 2010;239(2):703–14.

33. Paraoanu LE, et al. Expression patterns of neurexin-1 and neuroligins in brain and retina of the chick embryo: Neuroligin-3 is absent in retina.. Neurosci. Lett. 2006;395(2):114–7.

34. Varoqueaux F, et al. Neuroligins determine synapse maturation and function.. Neuron 2006;51(6):741–54.

35. Varghese M, et al. Autism spectrum disorder: neuropathology and animal models.. Acta Neuropathol. 2017;134(4):537–566.

36. Al-Jawahiri R, et al. SOX11 variants cause a neurodevelopmental disorder with infrequent ocular malformations and hypogonadotropic hypogonadism and with distinct DNA methylation profile. Genet. Med. 2022;3600(22):1–13.

37. Pelizzola M, et al. AMDA: An R package for the automated microarray data analysis. BMC Bioinformatics 2006;7:1–9.

38. Smyth GK. Linear Models and Empirical Bayes Methods for Assessing Differential Expression in Microarray Experiments. Stat. Appl. Genet. Mol. Biol. 2004;3(1):1–25.

39. Virtanen P, et al. SciPy 1.0--Fundamental Algorithms for Scientific Computing in Python. arXiv e-prints 2019;arXiv:1907.10121.

40. Pedregosa F, et al. Scikit-learn: Machine Learning in {P}ython. J. Mach. Learn. Res. 2011;12:2825–2830.

41. Hunter JD. Matplotlib: A 2D Graphics Environment. Comput. Sci. Eng. 2007;9(3):90– 95.

42. Waskom M, et al. seaborn doi:10.5281/zenodo.592845

43. Yang Y, et al. Molecular Findings Among Patients Referred for Clinical Whole-Exome Sequencing. JAMA 2014;312(18):1870.

44. Nambot S, et al. Clinical whole-exome sequencing for the diagnosis of rare disorders with congenital anomalies and/or intellectual disability: substantial interest of prospective annual reanalysis. Genet. Med. 2018;20(6):645–654.

45. Li H, Durbin R. Fast and accurate short read alignment with Burrows-Wheeler transform. Bioinformatics 2009;25(14):1754–1760.

46. Cingolani P, et al. A program for annotating and predicting the effects of single nucleotide polymorphisms, SnpEff. Fly (Austin). 2012;6(2):80–92.

47. Cannarella R, et al. Anti-Müllerian Hormone, Growth Hormone, and Insulin-Like Growth Factor 1 Modulate the Migratory and Secretory Patterns of GnRH Neurons. Int. J. Mol. Sci. 2021;22(5):2445.

48. Busnelli M, et al. Fat-Shaped Microbiota Affects Lipid Metabolism, Liver Steatosis, and Intestinal Homeostasis in Mice Fed a Low-Protein Diet. Mol. Nutr. Food Res. 2020;64(15):e1900835.

49. Budreck EC, Scheiffele P. Neuroligin-3 is a neuronal adhesion protein at GABAergic and glutamatergic synapses.. Eur. J. Neurosci. 2007;26(7):1738–48.

50. Cariboni A, et al. Dysfunctional SEMA3E signaling underlies gonadotropin-releasing hormone neuron deficiency in Kallmann syndrome.. J. Clin. Invest. 2015;125(6):2413–28.

