## Supplemental Data 1 for "Combined omic analyses reveal novel loss-of-function *NLGN3* variants in GnRH deficiency and autism"

### Supplemental Tables

#### Supplemental Table 1.

| Term ID | Term description | Observed gene count | Background gene count | FDR |
|---|---|---|---|---|
| E14 | | | | |
| rno04110 | Cell cycle | 26 | 123 | 4.03E-11 |
| rno04390 | Hippo signaling pathway | 18 | 149 | 0.00022 |
| rno03030 | DNA replication | 9 | 35 | 0.00026 |
| rno04550 | Signaling pathways regulating pluripotency of stem cells | 16 | 132 | 0.00031 |
| rno04350 | TGF-beta signaling pathway | 12 | 83 | 0.00077 |
| rno04520 | Adherens junction | 9 | 69 | 0.0084 |
| E17 | | | | |
| rno04060 | Cytokine-cytokine receptor interaction | 28 | 217 | 1.11E-08 |
| rno04062 | Chemokine signaling pathway | 21 | 168 | 1.53E-06 |
| rno04514 | Cell adhesion molecules (CAMs) | 15 | 156 | 0.00071 |
| E20 | | | | |
| rno04360 | Axon guidance | 25 | 171 | 1.31E-06 |
| rno04514 | Cell adhesion molecules (CAMs) | 22 | 156 | 1.01E-05 |

**Supplemental table 1.** List of selected KEGG pathways (FDR < 0.01)

**Supplemental table 2.**

| Term ID | Term description | Observed gene count | Background gene count | FDR |
|---|---|---|---|---|
| E14 | | | | |
| GO:0030335 | positive regulation of cell migration | 21 | 240 | 0.00082 |
| GO:0030334 | regulation of cell migration | 27 | 366 | 0.001 |
| GO:0000226 | microtubule cytoskeleton organization | 15 | 143 | 0.0015 |
| GO:0007010 | cytoskeleton organization | 27 | 391 | 0.0022 |
| GO:0007017 | microtubule-based process | 19 | 231 | 0.0028 |
| GO:0040011 | locomotion | 26 | 404 | 0.006 |
| E17 | | | | |
| GO:0060326 | cell chemotaxis | 21 | 76 | 4.50E-11 |
| GO:0006935 | chemotaxis | 27 | 172 | 1.49E-09 |
| GO:0022407 | regulation of cell-cell adhesion | 24 | 142 | 4.00E-09 |
| GO:0016477 | cell migration | 32 | 293 | 5.11E-08 |
| GO:0030155 | regulation of cell adhesion | 29 | 248 | 7.32E-08 |
| GO:0022409 | positive regulation of cell-cell adhesion | 17 | 87 | 2.22E-07 |
| GO:0040012 | regulation of locomotion | 37 | 419 | 3.78E-07 |
| GO:0030334 | regulation of cell migration | 34 | 366 | 4.52E-07 |
| GO:0045785 | positive regulation of cell adhesion | 21 | 151 | 6.26E-07 |
| GO:0040011 | locomotion | 34 | 404 | 3.12E-06 |
| GO:0070098 | chemokine-mediated signaling pathway | 10 | 30 | 3.16E-06 |
| GO:0071345 | cellular response to cytokine stimulus | 30 | 331 | 3.89E-06 |
| GO:0030335 | positive regulation of cell migration | 25 | 240 | 3.91E-06 |
| GO:0050920 | regulation of chemotaxis | 14 | 91 | 2.97E-05 |

| | | | | |
|---|---|---|---|---|
| GO:0050921 | positive regulation of chemotaxis | 12 | 68 | 4.46E-05 |
| GO:0022408 | negative regulation of cell-cell adhesion | 9 | 64 | 0.0022 |
| E20 | | | | |
| GO:0031344 | regulation of cell projection organization | 68 | 390 | 6.99E-22 |
| GO:0010975 | regulation of neuron projection development | 58 | 318 | 1.74E-19 |
| GO:0031346 | positive regulation of cell projection organization | 48 | 243 | 2.56E-17 |
| GO:0031175 | neuron projection development | 52 | 307 | 2.62E-16 |
| GO:0010976 | positive regulation of neuron projection development | 41 | 205 | 5.63E-15 |
| GO:0048812 | neuron projection morphogenesis | 37 | 179 | 6.86E-14 |
| GO:0007155 | cell adhesion | 48 | 309 | 8.35E-14 |
| GO:0061564 | axon development | 33 | 142 | 1.21E-13 |
| GO:0098609 | cell-cell adhesion | 28 | 130 | 6.76E-11 |
| GO:0007409 | axonogenesis | 26 | 112 | 9.52E-11 |
| GO:0098742 | cell-cell adhesion via plasma-membrane adhesion molecules | 21 | 75 | 5.49E-10 |
| GO:0040011 | locomotion | 46 | 404 | 3.59E-09 |
| GO:0050770 | regulation of axonogenesis | 20 | 92 | 5.51E-08 |
| GO:0031345 | negative regulation of cell projection organization | 20 | 110 | 6.25E-07 |
| GO:0010977 | negative regulation of neuron projection development | 17 | 93 | 5.30E-06 |
| GO:0016477 | cell migration | 31 | 293 | 8.87E-06 |
| GO:0050772 | positive regulation of axonogenesis | 12 | 45 | 8.97E-06 |

| | | | | |
|---|---|---|---|---|
| GO:0051493 | regulation of cytoskeleton organization | 27 | 233 | 9.54E-06 |
| GO:0007411 | axon guidance | 14 | 65 | 9.68E-06 |
| GO:0042330 | taxis | 22 | 173 | 2.39E-05 |
| GO:0048870 | cell motility | 32 | 328 | 2.61E-05 |
| GO:0007156 | homophilic cell adhesion via plasma membrane adhesion molecules | 11 | 42 | 2.74E-05 |
| GO:0001764 | neuron migration | 12 | 53 | 3.38E-05 |
| GO:0006935 | chemotaxis | 21 | 172 | 6.52E-05 |
| GO:0030516 | regulation of axon extension | 11 | 55 | 0.00021 |
| GO:0032886 | regulation of microtubule-based process | 14 | 97 | 0.0004 |
| GO:0007157 | heterophilic cell-cell adhesion via plasma membrane cell adhesion molecules | 7 | 23 | 0.00068 |
| GO:0051494 | negative regulation of cytoskeleton organization | 11 | 65 | 0.00071 |
| GO:0032970 | regulation of actin filament-based process | 18 | 171 | 0.0013 |
| GO:0040012 | regulation of locomotion | 32 | 419 | 0.0015 |
| GO:0007010 | cytoskeleton organization | 30 | 391 | 0.0021 |
| GO:0007017 | microtubule-based process | 21 | 231 | 0.0023 |
| GO:0007158 | neuron cell-cell adhesion | 4 | 6 | 0.0024 |
| GO:0045773 | positive regulation of axon extension | 7 | 30 | 0.0024 |
| GO:0016338 | calcium-independent cell-cell adhesion via plasma membrane cell-adhesion molecules | 4 | 7 | 0.0034 |
| GO:0007413 | axonal fasciculation | 5 | 15 | 0.0042 |
| GO:0070507 | regulation of microtubule cytoskeleton organization | 11 | 85 | 0.0044 |

| | | | | |
|---|---|---|---|---|
| GO:0030334 | regulation of cell migration | 27 | 366 | 0.0061 |
| GO:0050771 | negative regulation of axonogenesis | 6 | 29 | 0.0089 |
| GO:0031109 | microtubule polymerization or depolymerization | 5 | 19 | 0.0091 |
| GO:0032956 | regulation of actin cytoskeleton organization | 15 | 147 | 0.0049 |
| GO:0040013 | negative regulation of locomotion | 14 | 124 | 0.0032 |

**Supplemental table 2.** List of selected GO Biological Processes (FDR < 0.01)

**Supplemental table 3.**

| **HH/KS genes** | |
|---|---|
| *ANOS1* | *NSMF* |
| *CCDC141* | *NR0B1* |
| *CHD7* | *NTN1* |
| *DCC* | *OTUD4* |
| *DMXL2* | *PCSK1* |
| *FEZF1* | *PLXNA1* |
| *FGF17* | *PNPLA6* |
| *FGF8* | *POLR3A* |
| *FGFR1* | *POLR3B* |
| *FSHB* | *PROK2* |
| *GNRH1* | *PROKR2* |
| *GNRHR* | *RNF216* |
| *HS6ST1* | *SEMA3A* |
| *IGSF10* | *SEMA3E* |
| *IL17RD* | *SMCHD1* |
| *KISS1R* | *SOX10* |
| *KISS1R* | *SOX2* |
| *KLB* | *TAC3* |
| *LEP* | *TACR3* |
| *LEPR* | *TUBB3* |
| *LHB* | |

**Supplemental table 3.** List of known HH/KS causative genes used as 'input' genes to instruct ToppGene software.

### Supplemental Figures

### Supplemental Figure 1.

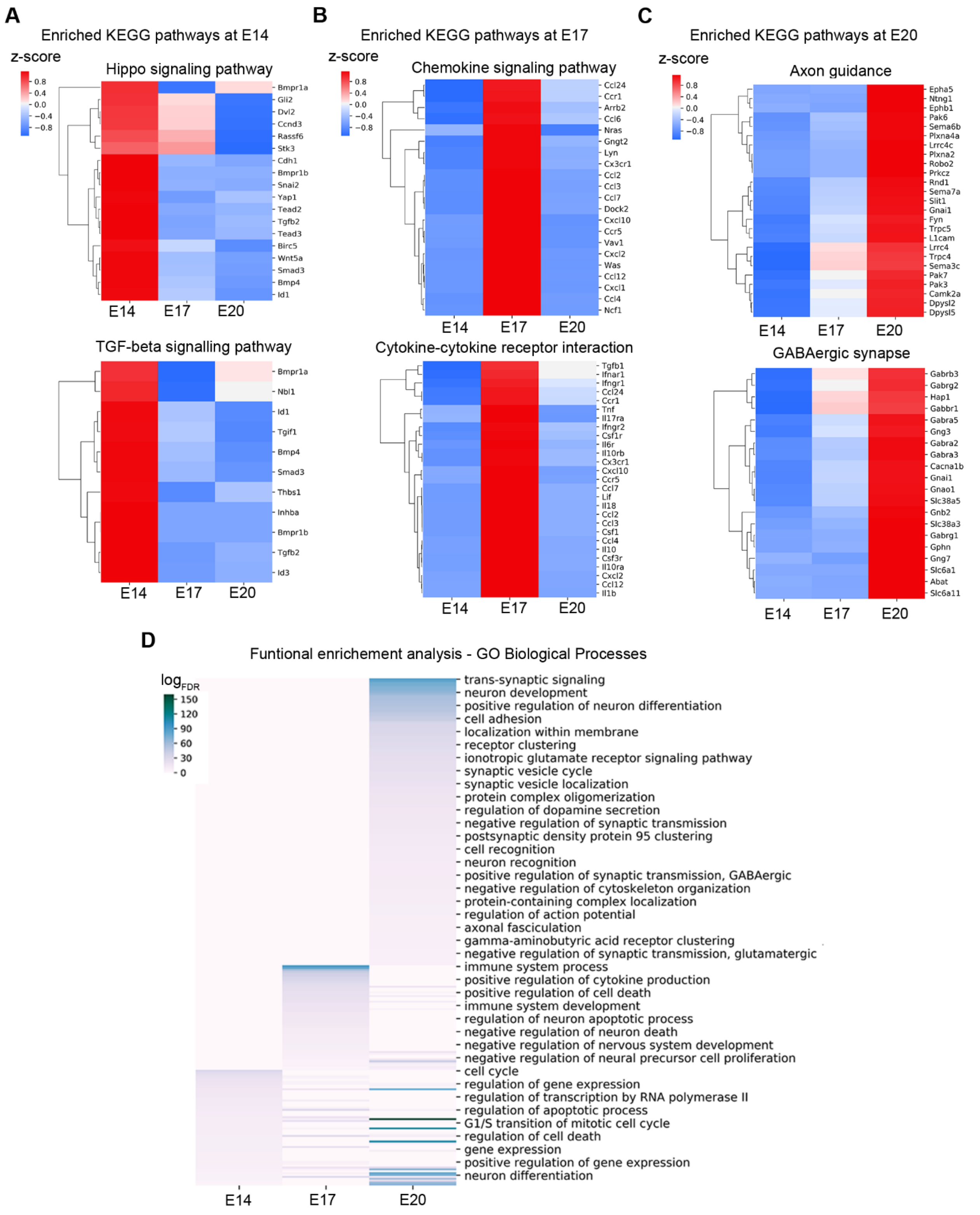

**Supplemental Figure 1. GFP$^{+}$ cells displayed specific gene expression signatures for each developmental stage.** A-C - Z-scored gene expression values for genes belonging to selected KEGG pathways are shown. Heat-maps representing examples of color-coded expression levels of genes belonging to significantly enriched KEGG pathways at E14 (A), E17 (B) and E20 (C). D - A functional enrichment analysis has been carried out for genes upregulated at E14, E17 and E20 timepoints and the enriched GO Biological Processes (FDR < 0.01) are summarized in the heatmap. Enrichment scores are reported as $\log_{FDR}$; higher values (deep blue) indicate highly enriched pathways, lower values (grey) indicate poorly enriched pathways.

**Supplemental Figure 2.**

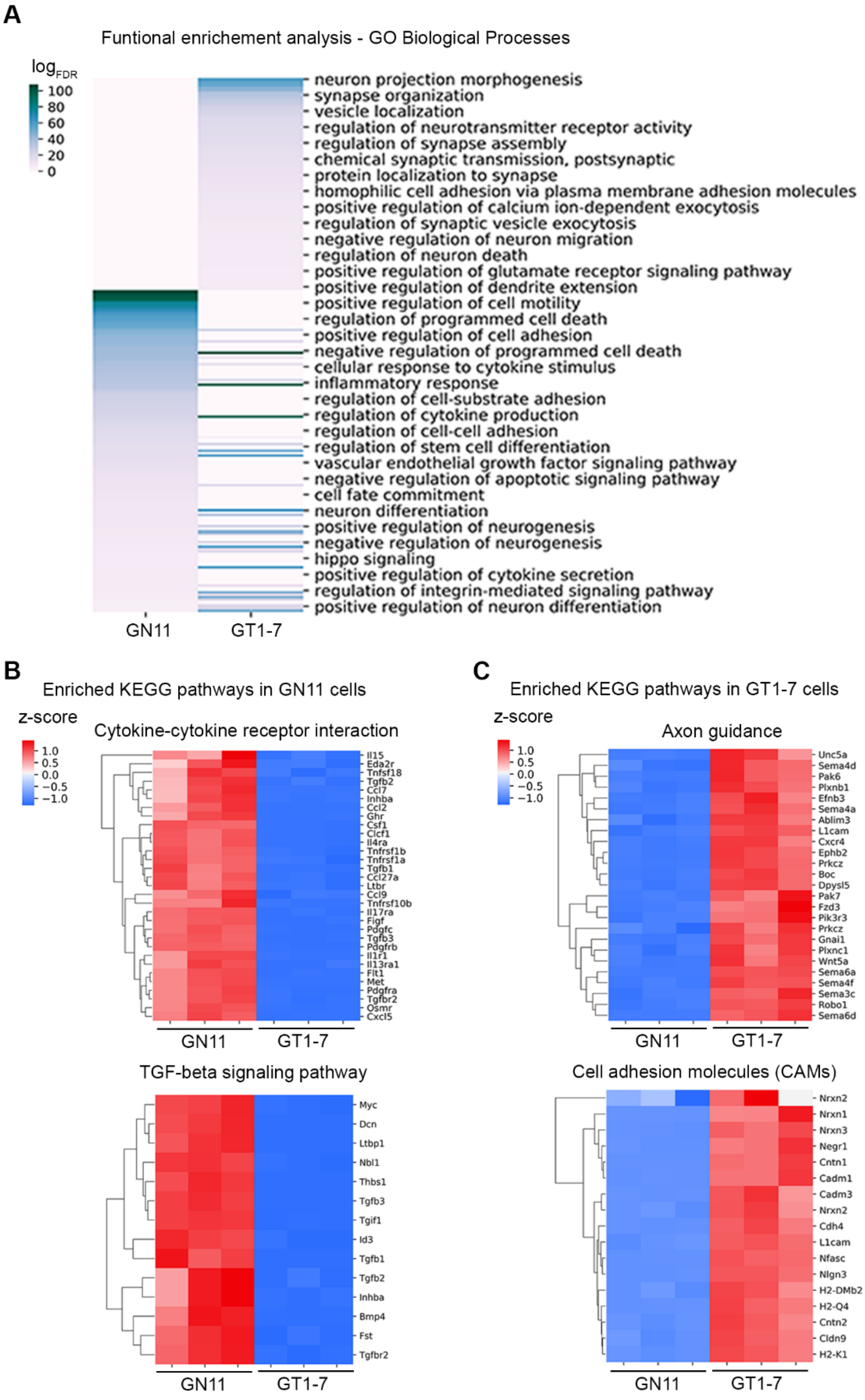

**Supplemental Figure 2. GN11 and GT1-7 cells displayed specific gene expression signatures.**

A - Enriched GO Biological Processes (F, FDR < $10^{-5}$) found by STRING functional enrichment analysis computed on DEGs between GN11 and GT1-7 cells. Enrichment scores are reported as $log_{FDR}$; higher values (deep blue) indicate highly enriched pathways, lower values (grey) indicate poorly enriched pathways. B,C - Z-scored gene expression values for genes belonging to selected KEGG pathways are shown. Heat-maps representing examples of color-coded expression levels of genes belonging to significantly enriched KEGG pathways at GN11 (B) and GT1-7 (C) cells.

**Supplemental Figure 3.**

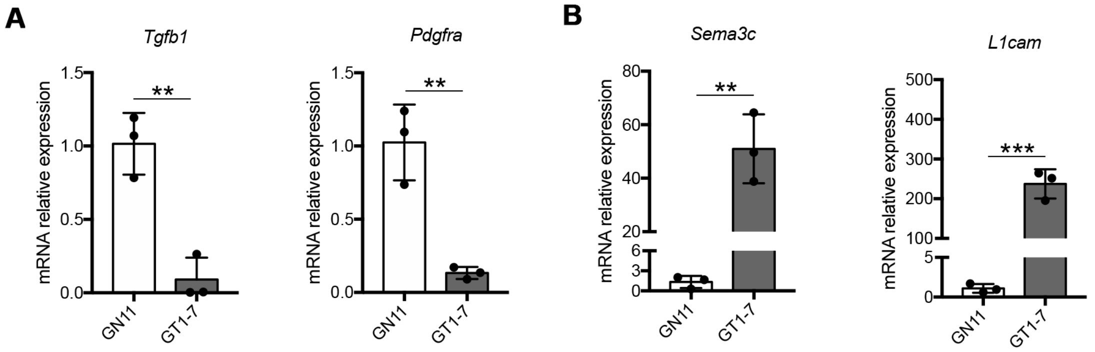


**Supplemental Figure 3. Validation of candidate genes by qPCR. A,B -** Expression levels of 2 representative early (A) and late (B) genes from the top 20 candidate gene lists (Table 3) quantified by qPCR. *Pdgfra* and *Tgfb1* were significantly upregulated in GN11 cells, whereas *Sema3c* and *L1cam* were significantly upregulated in GT1-7 cells. Data are presented as mean ± SD of 3 biological replicates. P values indicate Student's t test (** $P < 0.01$, *** $P < 0.001$).
